# Twitter Interaction to Analyze Covid-19 Impact in Ghana, Africa from March to July

**DOI:** 10.1101/2020.08.25.20182048

**Authors:** Josimar E. Chire Saire, Kobby Panford-Quainoo

## Abstract

Present pandemic generated by Coronavirus Covid-19 has stopped the world, from Tourism, Business, Education and more. Actually, there are reported cases in every continent from America to Europe, from Europe to Australia. Africa is one continent in process of development with a variety of languages, limitations/problems and of course, abundance of culture. The aim of this paper is to analyze the impact in one country located in the middle-west, named Ghana. The data source for the analysis is Twitter(Social Network) and the analysis is using a Text Mining approach. Besides, a composition of graphics are presented considering Google search, daily reported cases. The study is exploratory to know what topics are most frequent in Great Accra Region. The conclusions are: Twitter is useful to get data for the analysis, and the interest of user from Ghana was high at the beginning but it was decreasing from April until July, probably the people is tired or adapting themselves to the actual situation.

## 1 Introduction

The global pandemic, COVID-19, formally known as the severe acute respiratory syndrome coronavirus 2 (SARS-COV-2) [1], was declared a pandemic and a global threat by the World Health Organization (WHO) on 11-03-2020 [2]. Since December 2019, when its outbreak was first announced in the city of Wuhan in China, the disease has spread very rapidly to many other countries. As of 11-03-2020, when the disease was declared a global pandemic, a total of 6,546 cases and 355 deaths had been recorded by WHO [3].

Ghana had its first imported case of COVID-19 in Accra, the nation’s capital, and the regional capital of the Greater Accra Region, on 12-03-2020 [4],[5]. The disease then rapidly spread across the other regions. As of 3-08-2020, 10:35 GMT+2, a total of 37,014 cases had been confirmed. The Greater Accra Region alone accounted for about 18,882, which is about 51% of the total cases in Ghana[6].

The coronavirus disease is caused by a pathogen that attacks the respiratory system of humans. Non-infected persons may contract the disease after physical contact with an infected person or contaminated surfaces and objects. The disease is characterized by fever, dry cough, and tiredness, which manifests within 14 days of infection. Other symptoms include diarrhea, sore throat, conjunctivitis, headache, and loss of taste and smell [7], [8]. To slow down and control the spread of the virus, The WHO defined some measures in the absence of an approved cure [9]. Many governments soon adopted and implemented these measures in their own countries. These measures include physical distancing between persons, wearing face masks, avoiding physical interactions like handshaking, and hugging, and lockdowns, etc. These measures have been communicated to individuals through various communication channels, including social media platforms like Twitter, Facebook, etc.

As a result, concerned individuals and public health officials are able to stay up to date with research findings, statistical and precautionary measures. Others use these platforms to share their own opinions and sentiments towards media reports or news[10]. A careful analysis of what individuals talk about will help satisfy digital epidemiological needs for efficient disease surveillance and case severity analysis. Healthcare providers or policymakers will then understand the state of the situation, thereby making it easy to reach out with available remedies[11]. In this work, we use data sourced from Twitter, which is the most used social network after Facebook and YouTube[12], by a section of the Ghanaian population in the Greater Accra Region.

We contribute by providing a data-based assessment of the COVID-19 situation and the population’s response. This will aid healthcare providers and educators to assess how individuals at the target location feel, respond and understand matters concerning the COVID-19 pandemic. Based on this information, healthcare needs and decisions that are more suited to the population can be made and implemented.

## 2 Proposal

The proposal to analyze the situation in the country of Ghana follows the next steps:

– Select keywords related to covid19 pandemic
– Set the parameters to collect related data
– Pre-processing text
– Visualization to support Analysis

### 2.1 Select relevant terms

The scope of the analysis is Ghana, and considering access to Internet and concentration of population. The region selected is Great Accra, the selected terms are:

– ‘covid’, ‘coronavirus’, ‘sars’

### 2.2 Build the Query and Collect Data

The collection process is through Twitter Search function, with the next parameters:

– date: 01-03-2020 to 15-07-2020
– terms: the chosen words mentioned in previous subsection
– geolocalization: 5.646129,-0.065209, see Fig. 1
– language: English
– radius: 25.3347 km

**Fig. 1.**
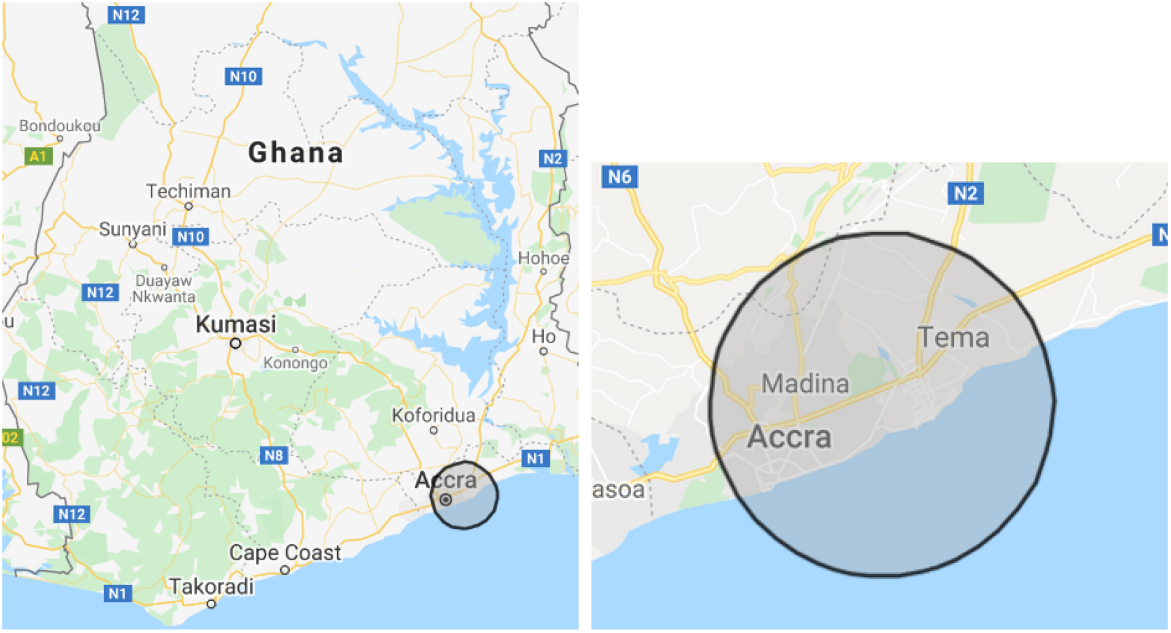
Geolocalization of Great Accra Region, Ghana

### 2.3 Preprocessing Data

This step is very important to take relevant words and this is the source to create graphics to help understanding the country.

– Uppercase to lowercase
– Eliminate alphanumeric symbols
– Delete stopwords, i.e. articles, pronouns, etc.
– Delete custom stopwords

### 2.4 Visualization

After cleaning the data, the text is ready to create graphics to help the understanding of the situation in Ghana during the last months.

– Bar plots to show the frequency of tweets per day, per month
– Cloud of words to visualize the most frequent terms per month
– Line plots to show the trend of data

## 3 Results

This section explains the results obtained and the respective analysis. First subsection 3.1 introduces the description of the dataset. Next subsections 3.2, 3.3 presents the frequency of post daily,monthly and 3.3, the most frequent terms per month. Finally, a crossing graphic is presented in 3.4 to analyze the relationship between Twitter, Google Trends, and new reported cases per day.

### 3.1 Dataset

This dataset is the result of the collection explained in 2.2 previously.

– Total size of dataset: 9475 tweets
– Fields: date(YYYY-MM-DD), text(alphanumeric)
– Range Date: 01-03-2020 to 15-07-2020
– Language: English

### 3.2 How is the frequency of post in Great Accra Region?

First, it is important to know how many tweets/posts related to the topic of the study because this can express the interest of the users about the topic around covid-19 pandemic. Figure 2, shows March, April were the months with highest number of posts and from April there is less tweets month after month.

**Fig. 2.**
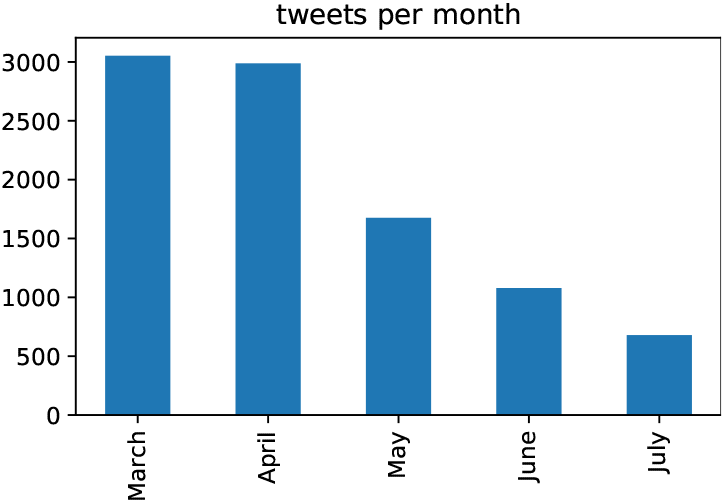
Number of Tweets per month

Splitting the previous graphic, it is possible to show the frequency daily. Then, figure 3 can summarize this information. Observing this graphic, from 01 to 27 March there is a constant increasing number of tweets but after 27 March, it starts to decrease. There are some peaks on March(27), April(13,19), May(19,31), June(14,24) and July(02), these numbers could be related to some events in the region.

The highlighted dates can be the start of an elaborated study around these days, then the study can be extended.

**Fig. 3.**
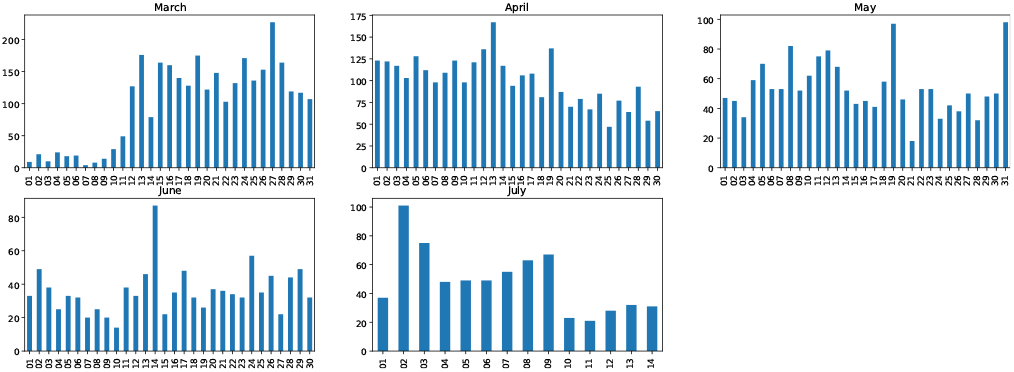
Number of Tweets per day

### 3.3 Relevant terms during the months?

The terms: covid19, coronavirus, sars were extracted because the intention is to know the most frequent terms around these keywords per month. First, graphic 4 shows case, people, pandemic, update, positive. Then, this keywords are more related to Public Health reports.

**Fig. 4.**
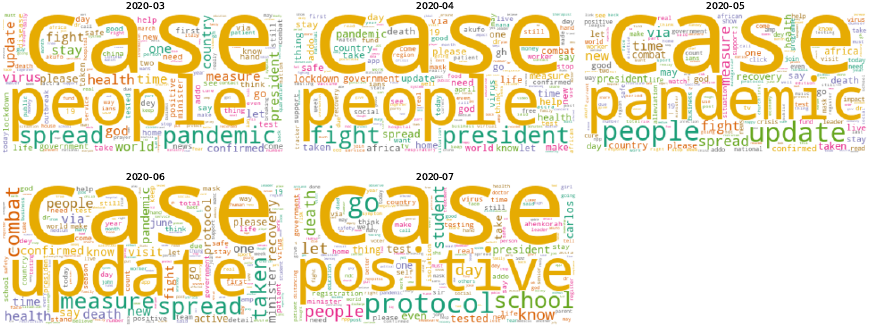
Cloud of words - Raw

Later, case and people terms are unconsidered and the result is graphic 5. Now, the most frequent are: pandemic, spread, health, president, update, protocol, school. Once more time, the posts are related to Public Health policies/actions of Ghana government.

**Fig. 5.**
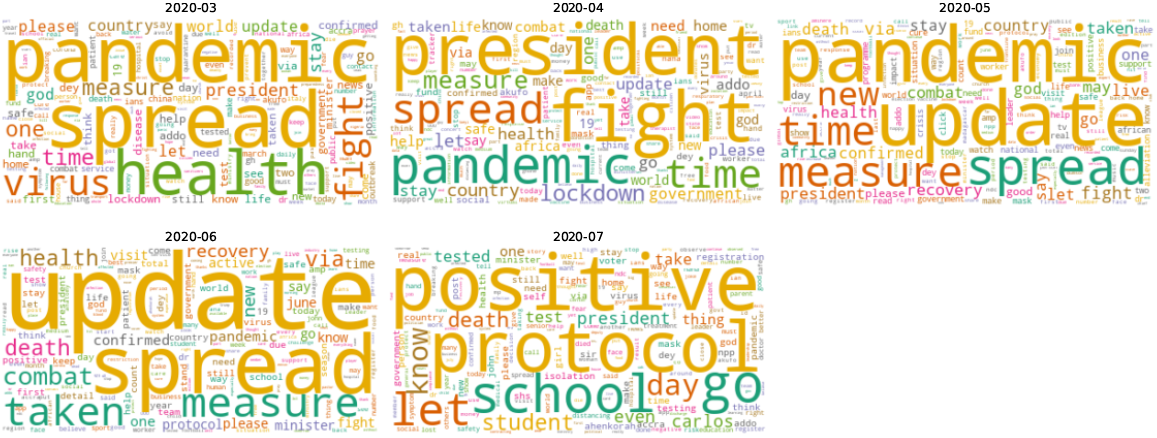
Number of Tweets per day

**Fig. 6.**
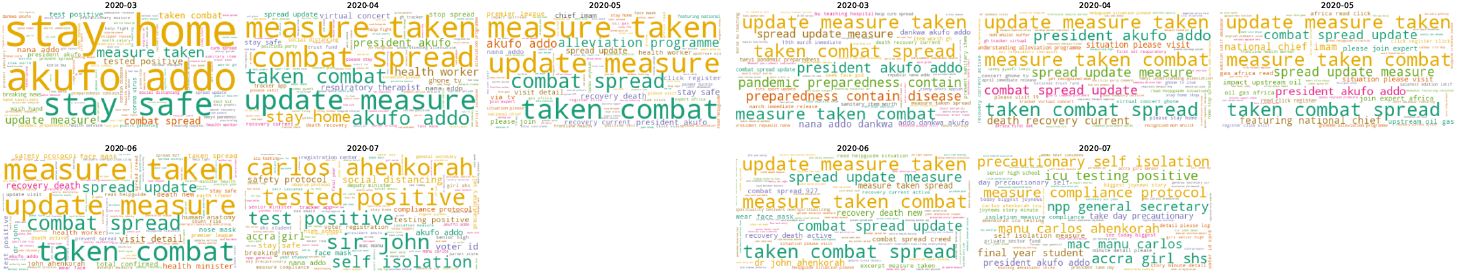
Number of Tweets per day

### 3.4 What is the relationship between Twitter Posts, Daily Reported Cases and Google Trends?

A valid question arises, how related can be posts of users with the actual pandemic? Therefore, graphic 7 presents the daily reported new cases, interest of users about Google search and Twitter posts.

**Fig. 7.**
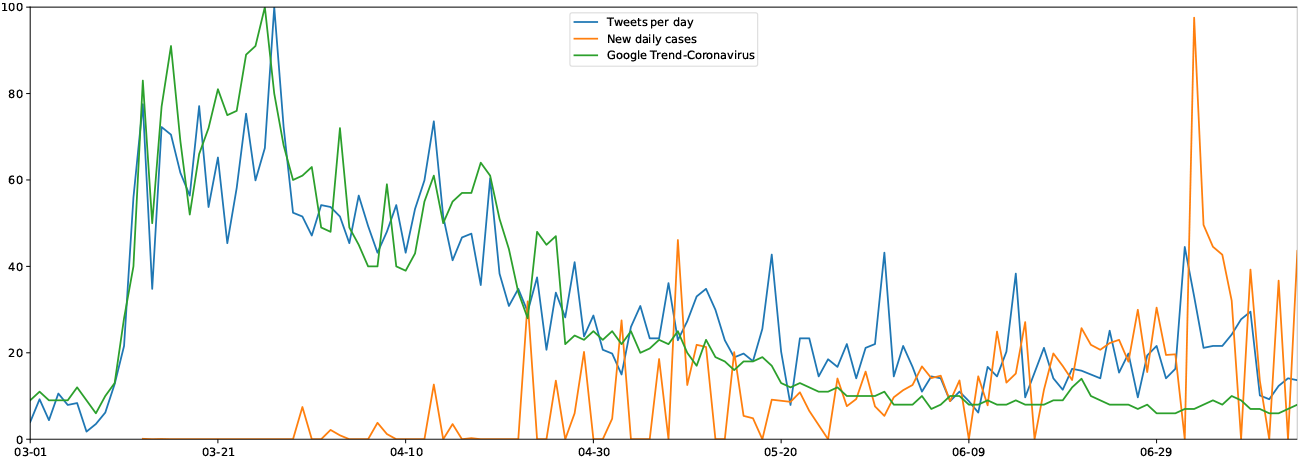
Graphic Daily reported cases, Google Search, Twitter Posts

The scale of Twitter posts was adapted to have values between 0-100 like Google trends. The data to elaborate graphic related to daily cases was downloaded from HDX website^3^.

There is visual correlation between searches about coronavirus topic and messasges posted daily. The interest of user has decreased during the time, it can be the result of many social, mental effects produced by lockdown or constant fear facing the pandemic. An opposite results with a increasing number of cases in the country, people can be tired of the context and started to adapt and reactivate their lives. This analysis is open for future work.

## 4 Conclusions

First, Twitter can be a useful source of data. Process to analyze text includes the cleaning of the text. Second, there was an increasing interest on the pandemic as topic in Ghana user at the beginning, during the first reported cases. But, during the time, the interest about the pandemic were decreasing. The evidence for this affirmation is found in Google searches, Twitter posts daily, in spite of the number of new cases still is growing up.

## Data Availability

Data can be required through emails of the authors.

## Footnotes

3 https://data.humdata.org/dataset/bc3589a6-04bc-4681-b531-7910ec800b4f

